# Comparison of a Target Trial Emulation Framework to Cox Regression to Estimate the Effect of Corticosteroids on COVID-19 Mortality

**DOI:** 10.1101/2022.05.27.22275037

**Authors:** Katherine L. Hoffman, Edward J. Schenck, Michael J. Satlin, William Whalen, Di Pan, Nicholas Williams, Iván Díaz

## Abstract

**Importance:** Communication and adoption of modern study design and analytical techniques is of high importance for the improvement of clinical research from observational data.

**Objective:** To compare (1) a modern method for causal inference including a target trial emulation framework and doubly robust estimation to (2) approaches common in the clinical literature such as Cox proportional hazards models. To do this, we estimate the effect of corticosteroids on mortality for moderate-to-severe coronavirus disease 2019 (COVID-19) patients. We use the World Health Organization’s (WHO) meta-analysis of corticosteroid randomized controlled trials (RCTs) as a benchmark.

**Design:** Retrospective cohort study using longitudinal electronic health record data for 28 days from time of hospitalization.

**Settings:** Multi-center New York City hospital system.

**Participants:** Adult patients hospitalized between March 1-May 15, 2020 with COVID-19 and not on corticosteroids for chronic use.

**Intervention:** Corticosteroid exposure defined as >0.5mg/kg methylprednisolone equivalent in a 24-hour period. For target trial emulation, interventions are (1) corticosteroids for six days if and when patient meets criteria for severe hypoxia and (2) no corticosteroids. For approaches common in clinical literature, treatment definitions used for variables in Cox regression models vary by study design (no time frame, one-, and five-days from time of severe hypoxia).

**Main outcome:** 28-day mortality from time of hospitalization.

**Results:** 3,298 patients (median age 65 (IQR 53-77), 60% male). 423 receive corticosteroids at any point during hospitalization, 699 die within 28 days of hospitalization. Target trial emulation estimates corticosteroids to reduce 28-day mortality from 32.2% (95% CI 30.9-33.5) to 25.7% (24.5-26.9). This estimate is qualitatively identical to the WHO’s RCT meta-analysis odds ratio of 0.66 (0.53-0.82)). Hazard ratios using methods comparable to current corticosteroid research range in size and direction from 0.50 (0.41-0.62) to 1.08 (0.80-1.47).

**Conclusion and Relevance:** Clinical research based on observational data can unveil true causal relationships; however, the correctness of these effect estimates requires designing the study and analyzing the data based on principles which are different from the current standard in clinical research.

**Key Points:** *Question:* How do modern methods for causal inference compare to approaches common in the clinical literature when estimating the effect of corticosteroids on mortality for moderate-to-severe coronavirus disease 2019 (COVID-19) patients?

*Findings:* In an analysis using retrospective data for 3,298 hospitalized COVID-19 patients, target trial emulation using a doubly robust estimation procedure successfully recovers a randomized controlled trial (RCT) meta-analysis benchmark. In contrast, analytic approaches common in the clinical research literature generally cannot recover the benchmark.

*Meaning:* Clinical research based on observational data can unveil true causal relations. However, the correctness of these effect estimates requires designing and analyzing the data based on principles which are different from the current standard in clinical research. Widespread communication and adoption of these analytical techniques are of high importance for the improvement of clinical research.

## Introduction

Observational databases are invaluable resources when randomized controlled trials (RCTs) are infeasible or unavailable. However, the correctness of the conclusions gleaned from analyses of observational data hinges on the careful consideration of study design principles and choice of estimation methodology.^1-4^

In this paper we contrast the use of target trial emulation using contemporary causal inference methods with various traditional analytical approaches using Cox regression. While most epidemiologists and statisticians agree on the importance of a well-defined exposure, outcome, and population of interest, the two strategies we compare differ significantly in the subsequent steps to choose a research question and data analysis method.

In the traditional approach to clinical research, the analysis proceeds by postulating a regression model according to the type of data available. For example, when faced with a time-to-event outcome, researchers automatically fit a Cox regression model (often due to limitations in knowledge, time, or software capabilities). The coefficients of the regression model are then used to answer to the clinical question of interest. We refer to this approach as a “model-first” approach, due to the primacy of the regression model.

A model-first approach induces multiple problems for the estimation of causal effects.^5^ First, regression coefficients often do not represent quantities of primary scientific interest or well-defined causal effects.^6^ Second, assumptions such as the proportional hazards assumption used in Cox models are rarely correct in medical research, since hazards cannot be proportional when a treatment effect changes over time.^7^ Third, regression models cannot correctly handle time-dependent feedback between confounders, treatment, and the outcome.^1^ Fourth, the model-first approach yields a tendency to interpret all coefficients in the model; a problem known as the Table 2 fallacy.^8^ Lastly, model-first approaches fail to account for the variance induced during model selection, thereby leading to incorrect statistical conclusions.^9^

Recent developments in the causal inference literature provide researchers with a number of tools to alleviate the aforementioned biases. Frameworks such as the target trial emulation^10^ and roadmap for causal inference^11^ allow researchers to proceed with a *question-first* approach. Instead of defaulting to effect measures provided by regression models, a question-first approach begins by defining a hypothetical target trial and subsequent target of inference that answers the scientific question of interest. This is the so-called estimand, or quantity to be estimated. After the estimand is chosen, researchers have the freedom to select an estimation technique which mitigates model misspecification biases. Incorporating these principles can help clarify the research question, determine study eligibility requirements, identify enrollment and follow-up times, decide whether sufficient confounder data are available, increase the likelihood of obtaining a correct estimate, and more.^12,13^

In this study, we compare a question-first approach against multiple model-first approaches for causal inference. Our case study is the effect of corticosteroids on mortality for moderate-to-severe COVID-19 patients using a retrospective cohort of patients at NewYork-Presbyterian Hospital (NYPH) during Spring 2020. Lack of guidance for clinical practice at the beginning of the pandemic meant that high variability existed in the administration and timing of corticosteroids (eFigure 1). Provider practice variability aids in the estimation of causal effects by yielding datasets with adequate natural experimentation, but the resulting complex longitudinal treatment patterns complicate study design and analytical methods. This observational dataset together with results from numerous RCTs on corticosteroids provides a unique opportunity to benchmark design and analysis methods. We benchmark our target trial emulation results against effect measures obtained in the World Health Organization (WHO)’s RCT meta-analysis.^14^

## Methods

This study was designed in April 2020, prior to the results of corticosteroid RCTs and resulting clinical guidance. It was approved by the Institutional Review Board at Weill Cornell Medicine with a waiver of informed consent (no. 20-04021909). This report follows the Strengthening the Reporting of Observational Studies in Epidemiology (STROBE) guidelines.^15^

### Hypothetical target trial

#### Question

What is the effect of a treatment regime of corticosteroids administered under the clinical indication of severe hypoxia on mortality for COVID-19 hospitalized patients?

#### Population

Inclusion criteria is adult COVID-19 positive patients who were admitted to NYPH’s Cornell, Lower Manhattan, or Queens locations. Cases are confirmed through reverse-transcriptase– polymerase chain-reaction assays performed on nasopharyngeal swab specimens. The tests are obtained upon hospital admission, i.e., at the same time of eligibility and time zero. Patients who have chronic use of corticosteroids prior to hospitalization or who are transferred into NYPH from an outside hospital are excluded.

#### Hypothetical treatment regime

Patients would be randomized on their first day of hospitalization to receive either (1)standard of care therapy (without corticosteroids) or (2)standard of care plus a corticosteroid regimen to be administered if and when criteria for severe hypoxia are met. The corticosteroid dosage is a minimum of 0.5 mg/kg body weight of methylprednisolone equivalent per 24-hour period and the duration of therapy is six days.^16^ Corticosteroids include prednisone, prednisolone, methylprednisolone, hydrocortisone, and dexamethasone and choice of drug is at the attending physician’s discretion. Severe hypoxia is defined as the initiation of high-flow nasal cannula, venti-mask, noninvasive or invasive mechanical ventilation, or an oxygen saturation of *<*93% after the patient is on 6 Liters of supplemental oxygen via nasal cannula.

#### Outcome and estimand

The primary outcome would be 28-day mortality from time of randomization. The contrast of interest is the 28-day mortality rate difference comparing actual receipt of the two treatment regimes (i.e., the per-protocol effect).

#### Data analysis plan

A hypothetical trial can assume no loss-to-follow-up. Under perfect compliance we would analyze the difference in proportion of patients who experienced the outcome between the two treatment regimes.

### Emulation using observational data

#### Data source and cohort

The target trial emulation uses retrospective data from patients who meet the hypothetical trial’s eligibility criteria March 1-May 15, 2020. Demographic, comorbidity, and outcome data were manually abstracted by trained medical professionals into a secure REDCap database.^17^ These were supplemented with an internal COVID data repository housing laboratory, procedure, medication, and flowsheet data documented during standard care.^18^ Patients are followed for 28 days from hospitalization and lost to follow-up by discharge or transfer to an external hospital system.

#### Treatment regimes and measurement

To emulate the target trial corticosteroid treatment regime, we estimate the effect of a hypothetical dynamic treatment regime,^19^ whereby each patient is administered six days of corticosteroids if and when they meet severe hypoxia criteria. This dynamic regime is contrasted with a static regime where patients never receive corticosteroids.

We measure severe hypoxia using vital signs and flowsheet data and define it in the same way as our target trial. We measure corticosteroid exposure using the medication administration record. We compute cumulative mg/kg dosing of corticosteroids over rolling 24-hour windows, and if a patient received *>*0.5 mg/kg methylprednisolone equivalent, they are denoted as having corticosteroids exposure that day.

Since patients in the observed data are subject to loss-to-follow-up, emulating the trial with observational data requires conceptualizing a hypothetical world where all patients are observed through 28 days. Effects in this hypothetical world can be estimated using observed patient data under assumptions articulated in the *Data Analysis* section. An illustration of the treatment regimes as they relate to the observed data are shown in Figure 1.

**Figure 1.**
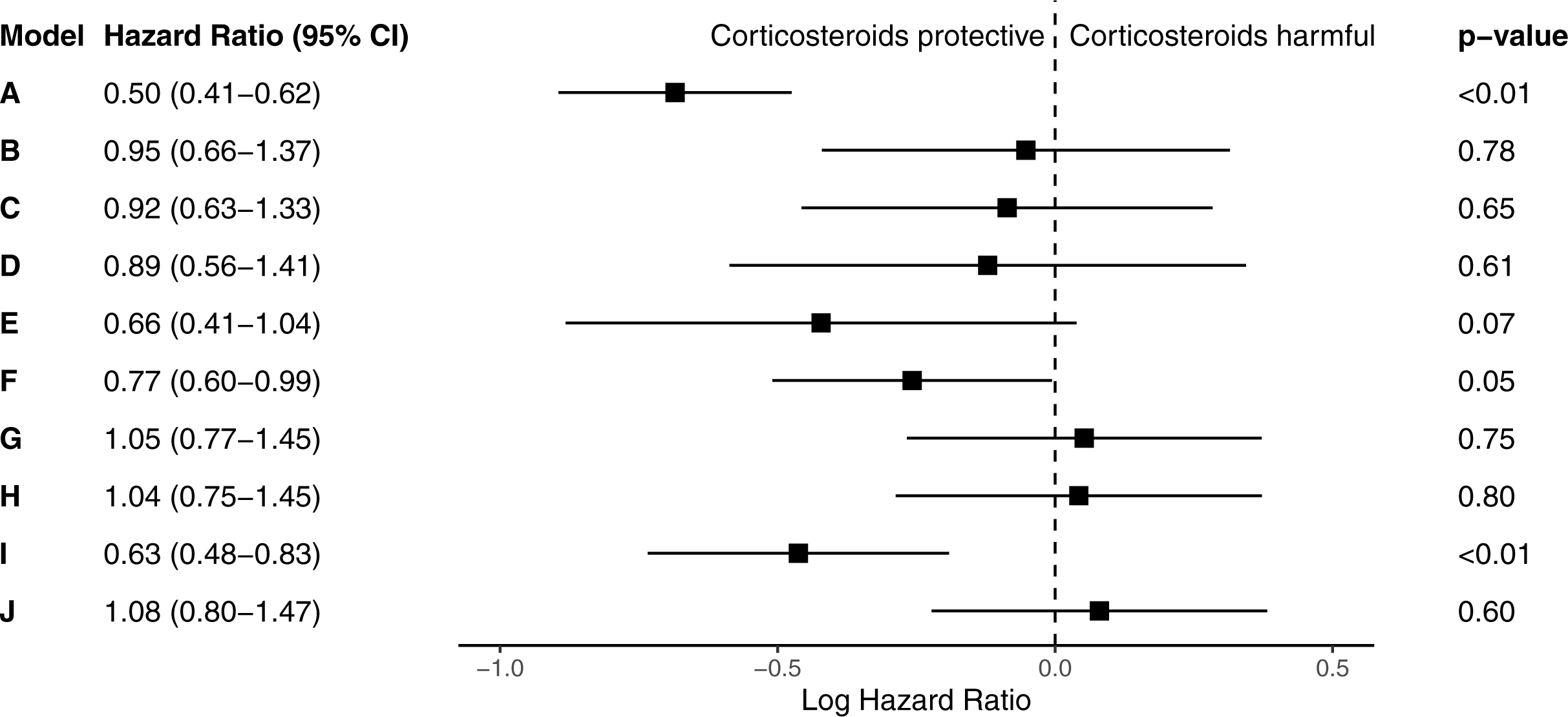
Illustrated example of two patients under the two hypothetical treatment regimes of our target trial emulation. Patient A reaches severe hypoxia criteria at study day 2 and is followed the entire study duration. Patient B never reaches severe hypoxia criteria and is lost to follow up after five study days. Under the dynamic corticosteroids regime (Intervention #1), Patient A receives 6 days of corticosteroids, and under Intervention #2 they receive no corticosteroids. Patient B does not receive corticosteroids under either treatment regime, however, in both hypothetical worlds they are observed for the entire study duration.

#### Confounding

In contrast to the hypothetical trial, treatment assignment in the observational study is not randomized and depends on physiological characteristics of each patient. We address confounding in our emulation by adjustment for confounders during data analysis. A set of confounders deemed sufficient for adjustment was determined through the expertise of a team of pulmonologists, intensivists, and microbiologists.

Baseline confounders include socio-demographics, Body Mass Index (BMI), comorbidities, and hospital admission location. Time-dependent confounders include vital signs, laboratory results, co-treatments, and mode of respiratory support. The measurement process (i.e., whether a clinician decided to measure these variables) is also an important confounder included in the analysis. Details of confounders are provided in eMethods. Figure 2 summarizes the relationship between confounders, treatment, and outcomes in the form of a Directed Acyclic Graph.

**Figure 2.**
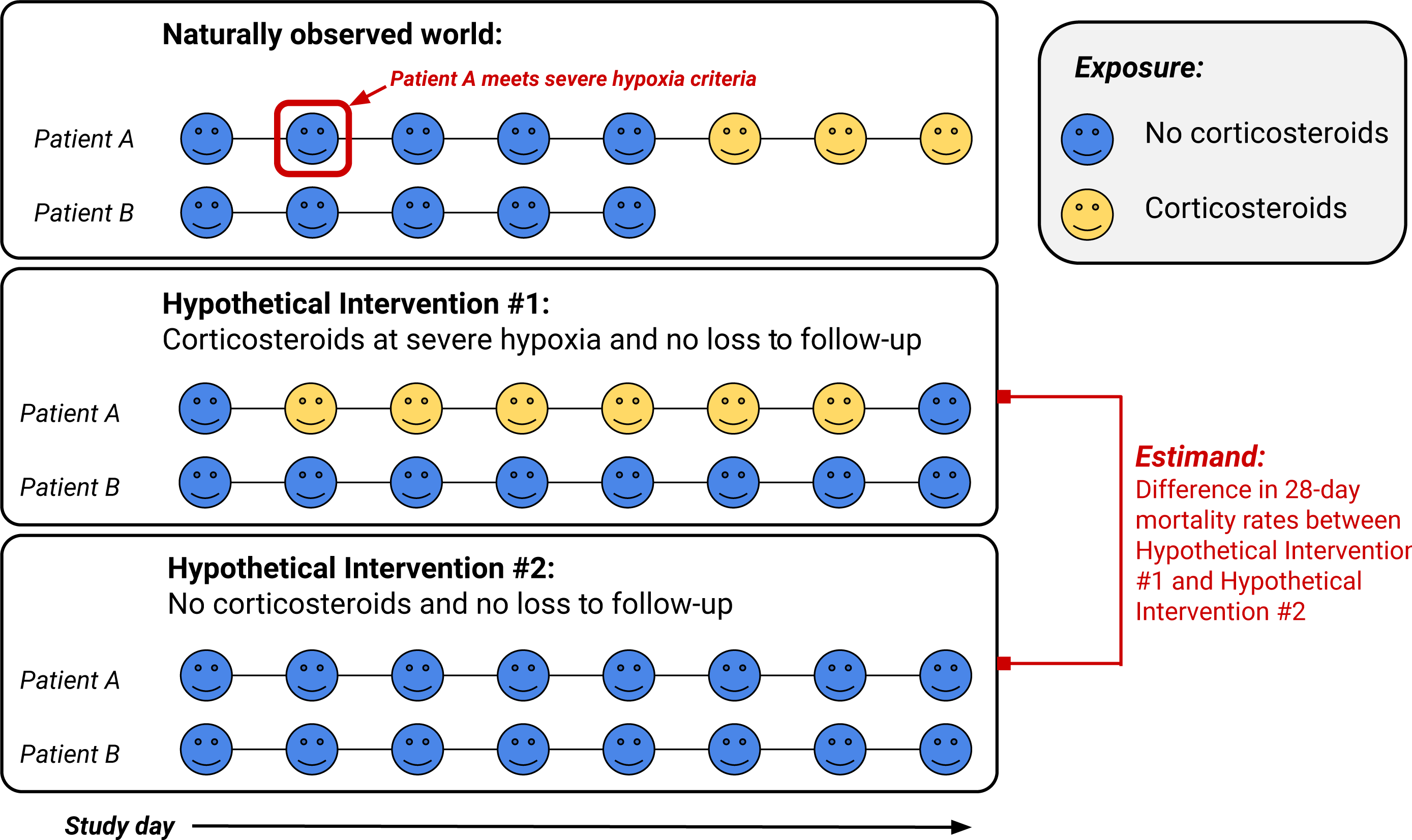
Illustrative Directed Acyclic Graph (DAG) showing the relationship between confounders *L*_*t*_, corticosteroid exposure *A*_*t*_, and mortality *Y*_*t*_. Baseline confounders are included in *L*_0_. For simplicity, loss-to-follow-up nodes are not shown. Abbreviations: BMI = Body Mass Index, BUN = Blood Urea Nitrogen, ACE/ARBs = Angiotensin-converting enzyme and Angiotensin receptor blockers.

#### Outcome and estimand

Our estimand of interest is the difference in 28-day mortality rates in a hypothetical world where we had implemented the two different corticosteroid treatment regimes, as well as an intervention to prevent loss-to-follow-up. Under the assumption that treatment and loss-to-follow-up each day are randomized conditional on the baseline and time-dependent confounders, this estimand is identifiable by a longitudinal g-computation formula.^20^ It is important to emphasize that conditional randomization is a key assumption without which the target emulation may fail. This g-formula will be our estimand of interest, but we note that it is not the only possible identification strategy (see eMethods).

#### Data analysis plan

When using the g-formula to identify causal effects, correct emulation of a target trial requires proper adjustment for measured confounding. It is important to use estimation methods capable of fitting the data using flexible mathematical relationships so that confounding is appropriately removed, especially when the number of baseline and time-dependent confounders is large.

Methods to estimate the g-computation formula (e.g., inverse probability weighting (IPW), parametric g-formula, targeted minimum loss-based estimators (TMLE), sequentially doubly robust estimators (SDR), etc.)^21,22^ rely on two kinds of mathematical models: (i) the outcome as a function of the time-dependent confounders, and (ii) treatment as a function of time-dependent confounders. Methods that use only one of these models are often called *singly robust*, because their correctness relies on the ability to correctly specify one of the models (e.g., IPW relies on estimating treatment models correctly). Methods that use both of these models are often called *doubly robust*, because they remain correct under misspecification of one of the two models.

Furthermore, doubly robust estimators such as TMLE and SDR allow the use of machine learning to flexibly fit relevant treatment and outcome regressions.^23,24^ This is desirable because these regression functions might include complex relationships, and capturing those relationships is not possible using simpler regression such as the Cox model.^25^

The primary analysis is conducted using SDR estimation with a dynamic intervention, time-varying confounders, and a time-to-event outcome. An ensemble of machine learning models using the super learner algorithm is used to estimate the regressions for treatment and outcome.^26,27^ Additional details, including sensitivity analyses, an illustrated analytical file (eFigure 2), and code tutorial, are available in eMethods.

### Model-first approaches

For contrast with the target trial emulation strategy, we review methodology of papers cited in Chaharom et al.’s^28^ COVID-19 corticosteroids meta-analysis, and then analyze the data using study designs common in those papers. The data source and outcome are the same as the above target trial. Modifications to the cohort, confounders, and treatment definitions to accommodate the model-first approaches are outlined below.

#### Point-treatment Cox models

The first approach we explore is a regression for mortality with a point-treatment variable. The inclusion criteria and time zero are defined as the time of meeting hypoxia criteria, which is the intended indication for corticosteroids. A study design using this approach entails several choices, including defining a range of time relative to inclusion criteria for a patient to be considered “treated”. Once this range is determined, researchers must decide how to handle patients treated before the inclusion time begins or after the treatment interval ends, as well as those who experience the outcome within the treatment interval.

We fit Cox models using data sets obtained from various design choices, summarized in Table 1. Baseline confounders and time-dependent confounders from day zero are included as adjustment variables. The exponentiated coefficient for corticosteroids is interpreted as the hazard ratio for corticosteroid exposure within the defined treatment window for moderate-to-severe COVID-19 patients.

**Table 1:**
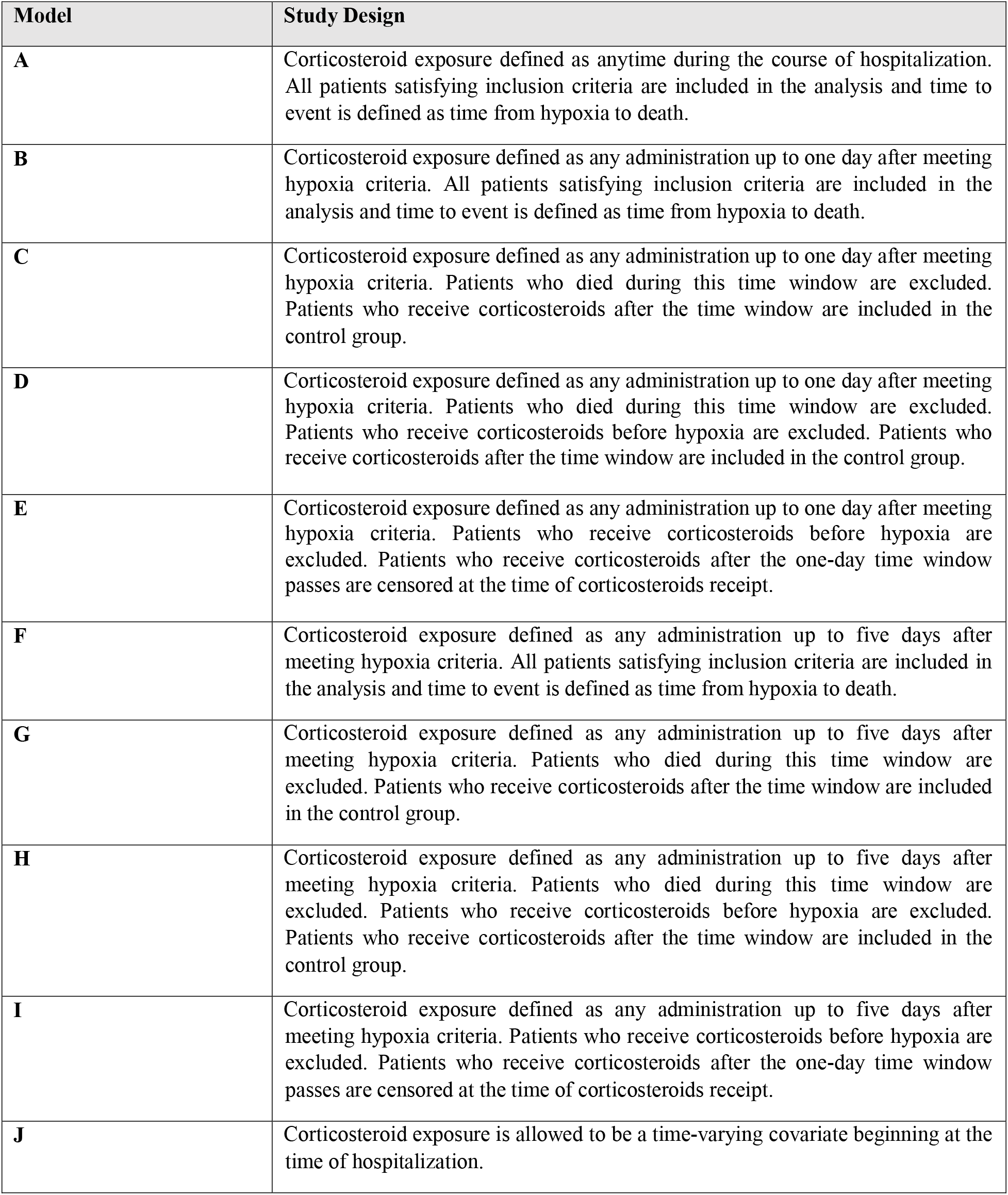
Study design specifications for the model-first approaches.

| Model | Study Design |
| --- | --- |
| **A** | Corticosteroid exposure defined as anytime during the course of hospitalization. All patients satisfying inclusion criteria are included in the analysis and time to event is defined as time from hypoxia to death. |
| **B** | Corticosteroid exposure defined as any administration up to one day after meeting hypoxia criteria. All patients satisfying inclusion criteria are included in the analysis and time to event is defined as time from hypoxia to death. |
| **C** | Corticosteroid exposure defined as any administration up to one day after meeting hypoxia criteria. Patients who died during this time window are excluded. Patients who receive corticosteroids after the time window are included in the control group. |
| **D** | Corticosteroid exposure defined as any administration up to one day after meeting hypoxia criteria. Patients who died during this time window are excluded. Patients who receive corticosteroids before hypoxia are excluded. Patients who receive corticosteroids after the time window are included in the control group. |
| **E** | Corticosteroid exposure defined as any administration up to one day after meeting hypoxia criteria. Patients who receive corticosteroids before hypoxia are excluded. Patients who receive corticosteroids after the one-day time window passes are censored at the time of corticosteroids receipt. |
| **F** | Corticosteroid exposure defined as any administration up to five days after meeting hypoxia criteria. All patients satisfying inclusion criteria are included in the analysis and time to event is defined as time from hypoxia to death. |
| **G** | Corticosteroid exposure defined as any administration up to five days after meeting hypoxia criteria. Patients who died during this time window are excluded. Patients who receive corticosteroids after the time window are included in the control group. |
| **H** | Corticosteroid exposure defined as any administration up to five days after meeting hypoxia criteria. Patients who died during this time window are excluded. Patients who receive corticosteroids before hypoxia are excluded. Patients who receive corticosteroids after the time window are included in the control group. |
| **I** | Corticosteroid exposure defined as any administration up to five days after meeting hypoxia criteria. Patients who receive corticosteroids before hypoxia are excluded. Patients who receive corticosteroids after the one-day time window passes are censored at the time of corticosteroids receipt. |
| **J** | Corticosteroid exposure is allowed to be a time-varying covariate beginning at the time of hospitalization. |

These point-treatment estimates apply only to the hypoxic population. They are different from the effects in the target trial emulation, which apply to the population of hospitalized patients. These effects are the closest possible analog we can obtain within a model-first framework using a point-treatment.

#### Time-varying Cox models

In the second model-first approach, we fit a time-varying Cox model for time to mortality up to 28 days from the day of hospitalization. This model uses the entire cohort and contains baseline and time-dependent confounders, as well as daily corticosteroid administration. The coefficient for corticosteroids is exponentiated and used as an estimate of the hazard ratio for corticosteroids on mortality in hospitalized COVID-19 patients.

### RCT benchmark

Several RCTs have established the effectiveness of corticosteroids in the treatment of moderate-to-severe COVID-19 patients.^29-31^ The WHO performed a meta-analysis of seven such RCTs and estimated the OR of mortality to be 0.66 (95% CI 0.53-0.82).^14^ We use this estimate, as well as supporting evidence from other RCT meta-analyses^28,32^ to benchmark our results. A discussion of assumptions for benchmarking, along with comparisons of our target trial study design, population, and treatment arms to the benchmark RCTs (eTables 1-3), is provided in the Appendix.

## Results

### Target trial emulation

In the target trial emulation analysis, all 3,298 patients who were admitted to the hospital are analyzed. Table 2 and eTable 4 display characteristics of the cohort, and eTable 5 describes the informative measurement process. There were 1,690 patients who reached severe hypoxia and 423 patients who received corticosteroids at any point during follow-up. 699 patients died before 28 days.

**Table 2.** Demographics and outcome for study cohort, overall and stratified by any corticosteroid exposure.

| Characteristic<sup>a</sup> | Overall<br>[N=3,298] | Corticosteroid<br>Never [N=2,875] | Corticosteroids<br>Ever [N=423] |
| --- | --- | --- | --- |
| **Age** | 65 (53, 77) | 65 (52, 77) | 67 (58, 75) |
| **Sex** |  |  |  |
| **Female** | 1,328 (40%) | 1,178 (41%) | 150 (35%) |
| **Male** | 1,970 (60%) | 1,697 (59%) | 273 (65%) |
| **Race<sup>b</sup>** |  |  |  |
| **Asian** | 602 (18%) | 517 (18%) | 85 (20%) |
| **Black** | 399 (12%) | 352 (12%) | 47 (11%) |
| **White** | 938 (28%) | 818 (28%) | 120 (28%) |
| **Other** | 1,141 (35%) | 1,009 (35%) | 132 (31%) |
| **Unknown or declined** | 218 (6.6%) | 179 (6.2%) | 39 (9.2%) |
| **Ethnicity** |  |  |  |
| **Hispanic or Latinx** | 1,117 (34%) | 994 (35%) | 123 (29%) |
| **Non-Hispanic or Latinx** | 1,585 (48%) | 1,388 (48%) | 197 (47%) |
| **Unknown or declined** | 596 (18%) | 493 (17%) | 103 (24%) |
| **BMI<sup>c</sup>** | 27 (23, 31) | 27 (23, 31) | 28 (24, 32) |
| **Home supplemental oxygen** | 312 (9.5%) | 286 (9.9%) | 26 (6.1%) |
| **Coronary Artery Disease** | 460 (14%) | 402 (14%) | 58 (14%) |
| **Diabetes Mellitus** | 1,033 (31%) | 891 (31%) | 142 (34%) |
| **Hypertension** | 1,780 (54%) | 1,544 (54%) | 236 (56%) |
| **Cerebral Vascular Event** | 225 (6.8%) | 193 (6.7%) | 32 (7.6%) |
| **Cirrhosis** | 35 (1.1%) | 30 (1.0%) | 5 (1.2%) |
| **CKD/ESRD** | 159 (4.8%) | 146 (5.1%) | 13 (3.1%) |
| **Asthma** | 180 (5.5%) | 145 (5.0%) | 35 (8.3%) |
| **COPD** | 134 (4.1%) | 100 (3.5%) | 34 (8.0%) |
| **Active cancer** | 136 (4.1%) | 118 (4.1%) | 18 (4.3%) |
| **Immunosuppressed** | 51 (1.5%) | 44 (1.5%) | 7 (1.7%) |
| **ILD** | 5 (0.2%) | 3 (0.1%) | 2 (0.5%) |
| **HIV** | 35 (1.1%) | 33 (1.1%) | 2 (0.5%) |
| **Active smoker** | 104 (3.2%) | 93 (3.2%) | 11 (2.6%) |
| **Former smoker** | 543 (16%) | 442 (15%) | 101 (24%) |
| ***Outcome: 28-day mortality*** | 699 (21%) | 574 (20%) | 125 (30%) |
| <sup>a</sup>All continuous variables are reported as median (interquartile range) and categorical variables are n (%). Abbreviations: BMI=Body Mass Index, CKD=Chronic Kidney Disease, ESRD=End Stage Renal Disease, COPD=Chronic Obstructive Pulmonary Disease, ILD=Interstitial Lung Disease, HIV=Human Immunodeficiency Virus. <sup>b</sup>Other race category includes American Indian or Alaskan Native, Pacific Islander, multiracial, or a patient response of “some other race”. <sup>c</sup>190 (5.8 %) patients did not have BMI data available. |  |  |  |

The estimated mortality rate under our no corticosteroids regime is 32.2% (95% CI 30.9-33.5). The estimated mortality rate under our corticosteroids regime is 25.7% (24.5-26.9). This yields an estimated mortality reduction of 6.5% (5.7-7.4) if this policy had been implemented. Sensitivity analyses (see Appendix) yield near-identical results.

### Model-first approaches

In the subset of patients who met severe hypoxia, 72 patients received corticosteroids within one day of hypoxia and 191 patients received corticosteroids within 5 days of hypoxia. There were 18 and 451 patients who died within one and five days of hypoxia without receiving corticosteroids, respectively.

Model A, which defined corticosteroid exposure as anytime during hospitalization, yielded an HR of 0.50 (0.41-0.62). Models B-I, which placed either a one- or five-day limit on corticosteroids treatment from the time of hypoxia, yielded mostly non-significant HRs in both directions (B: 0.95 (0.66-1.37), C: 0.92 (0.63-1.33), D: 0.89 (0.56-1.41), E: 0.66 (0.41-1.04), G: 1.05 (0.77-1.45), H: 1.04 (0.75-1.45)). The exception to this was Model I, which excluded patients who died before five days and estimated the HR to be 0.63 (0.48-0.83). Model F also reached statistical significance, 0.77 (0.60-0.99), and was the result of a 5-day treatment window with no exclusion or censoring variations. The time-varying Cox model yielded an HR of 1.08 (0.80-1.47). Figure 3 summarizes the model-first results.

**Figure 3.**
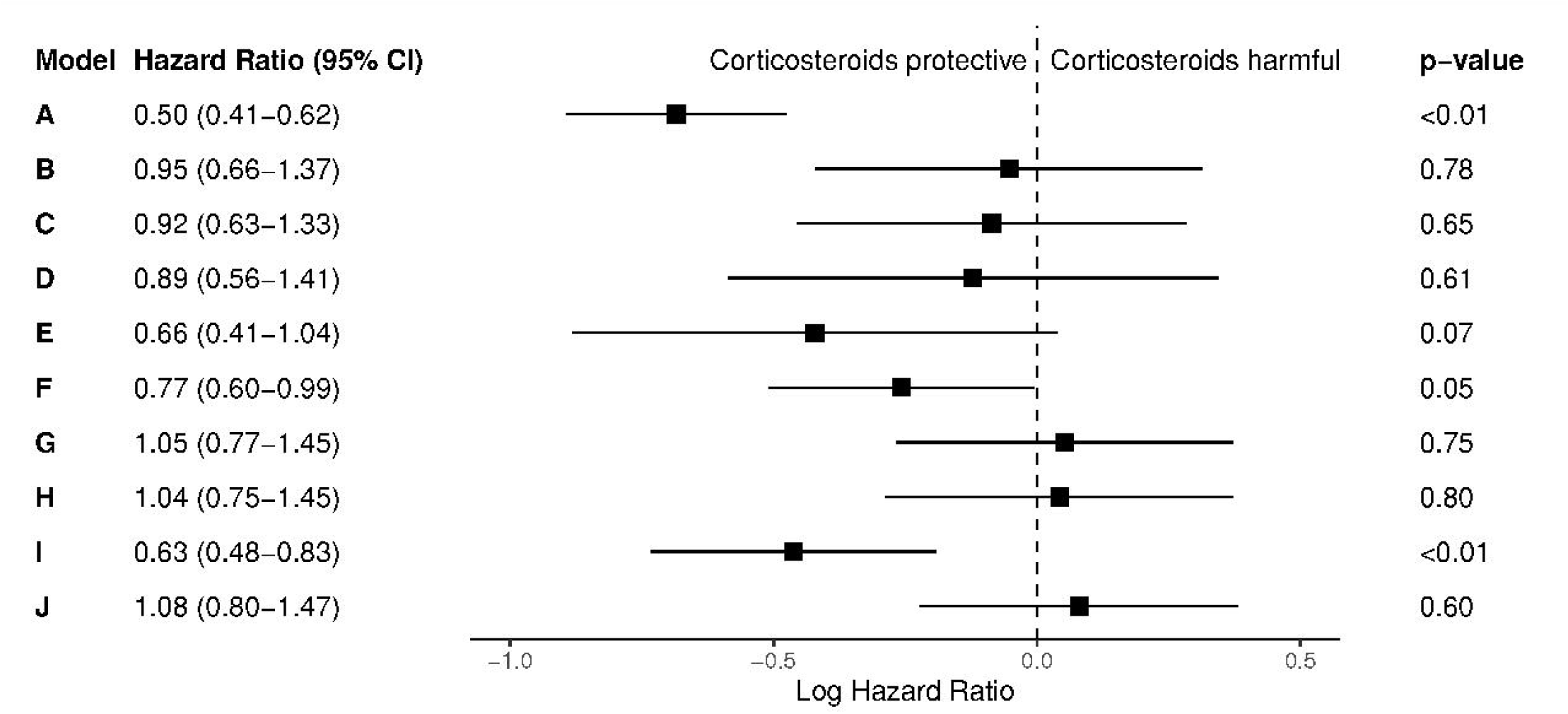
Forest plot of model-first results. Study designs A-J correspond to Table 1’s specifications.

## Discussion

Our research illustrates how a question-first approach can aid in devising an optimal design and choice of estimation procedure for an analysis of observational data. We show that using the target trial framework succeeds in recovering the benchmark causal effect obtained in RCTs. Our estimate that corticosteroids would reduce overall 28-day mortality in a hospitalized cohort is equivalent to an OR of 0.73 (0.68-.74), which is qualitatively identical to the WHO’s estimate of 0.66 (0.53-0.82). Our study design allowed us to conceptualize a meaningful intervention, i.e., randomize patients at hospitalization but do not give corticosteroids unless the patient becomes severely hypoxic. Our analysis plan enabled us to flexibly adjust for a large number of potential time-dependent confounders.

In contrast, the majority of the model-first approaches could not recover the RCT benchmark using the same data source. This finding aligns with other corticosteroids research; a recent meta-analysis containing observational analyses on over 18,000 patients found no overall effect for corticosteroids on mortality (OR 1.12, (0.83–1.50)).^28^ The task of creating reliable evidence from complex longitudinal data is not an easy one, and many of these studies suffer from flawed designs.

We found most studies in the current observational corticosteroids literature allowed the “treated” group to receive corticosteroids anytime during hospitalization.^33-35^ This is problematic because it introduces immortal time and biases results towards a protective effect of corticosteroids.^35^ A few studies did limit the treatment time frame in an effort to diminish immortal time bias. The “grace period” for treatment was handled in various ways, e.g. excluding patients who die prior to a time window after inclusion criteria,^37^ or excluding patients who receive treatment after the treatment window ends.^38,39^ Both exclusions may lead to bias and spurious associations.^1^ An alternative to exclusion is censoring patients at their time of receiving treatment if that time is after the treatment window passes, however, Cox regression cannot handle time-dependent censoring.^1^

In addition to these issues, it is often unclear in the current literature how patients who receive corticosteroids prior to meeting inclusion criteria are handled in the analysis.^33-35,40^ A related issue is that corticosteroids can affect severity of illness. All of the point-treatment studies are thus subject to collider bias by subsetting to severely ill patients.^41^ While the time-varying Cox approach does not suffer from the same time-alignment biases as the point-treatment design, it cannot properly account for time-dependent confounders.^1^ These biases appear in our model-first results; the study designs which result in a statistically significant protective effect of corticosteroids suffer from extreme immortal time bias through undefined or extended treatment time windows (A,F,I).

### Limitations

First, while the pre-RCT study time frame is ideal for natural experimentation and the estimation of causal effects, it includes surge conditions and rapidly changing clinical practice, challenging the assumptions needed for transportability and benchmarking. Second, we cannot rule out unmeasured confounding in the treatment, censoring, or outcome mechanisms. Specifically, the different discharge pathways (home, nursing home, etc.) may be associated to unmeasured patient characteristics and lead to very different outcomes. Third, we did not have the data to look at individual corticosteroid types, making comparisons to a specific RCT impossible. Fourth, the binning of our data into 24-hour intervals may induce issues related to the correct time-ordering of events (see Appendix).

### Conclusions

This study serves as an example in which the current standard for clinical research methods fails to recover the correct treatment effect where a modern causal inference method succeeds. Using observational data to guide clinical practice is possible but relies on the use of contemporary statistical and epidemiological principles. We hope this study and accompanying technical guide encourages adoption of similar innovative techniques into study designs and statistical analyses for observational medical research.

## Data Availability

Data produced in the present study are available in HIPAA compliant de-identified form upon reasonable request to the authors.

https://github.com/kathoffman/steroids-trial-emulation

## Acknowledgements

The authors thank all of the healthcare workers who courageously expanded their roles during the pandemic’s surge conditions. This work was made possible through data provided by the Cornell COVID-19 Registry, led by Parag G. Goyal, M.D., Justin Choi, M.D., Laura Pinheiro, Ph.D., and Monika Safford, M.D., of Weill Cornell Medicine. The authors would like to acknowledge the chart abstractors, which included a team of Weill Cornell Medicine medical students and NewYork-Presbyterian/Weill Cornell Medical Center house staff. The authors also thank the contributions to this work of the Architecture for Research Computing in Health team.

## Author contributions

Katherine L. Hoffman, M.S. had full access to all the data in the study and takes responsibility for the integrity of the data and the accuracy of the data analysis.

## References

[1] Robins JM Hernán MA. Causal Inference: What If. Boca Raton: Chapman & Hall; 2020.

[2] Miguel A. Hernán. Methods of public health research — strengthening causal inference from observational data. New England Journal of Medicine. 2021; 385(15):1345–1348. doi: 10.1056/NEJMp2113319. URL https://doi.org/10.1056/NEJMp2113319. PMID: 34596980.

[3] Steven S. Henley, Richard M. Golden, and T. Michael Kashner. Statistical modeling methods: challenges and strategies. Biostatistics & Epidemiology. 2020;4(1):105–139. doi: 10.1080/24709360.2019.1618653. URL https://doi.org/10.1080/24709360.2019.1618653.

[4] Mohammad Ali Mansournia, Mahyar Etminan, Goodarz Danaei, Jay S Kaufman, and Gary Collins. Handling time varying confounding in observational research. BMJ. 2017;359. ISSN 0959-8138. doi: 10.1136/bmj.j4587. URL https://www.bmj.com/content/359/bmj.j4587.

[5] Leo Breiman. Statistical modeling: The two cultures (with comments and a rejoinder by the author). Statistical science. 2001;16(3):199–231.

[6] Miguel A. Hernán. The Hazards of Hazard Ratios. Epidemiology. 2010;21(1):13–15, 01. doi: 10.1097/EDE.0b013e3181c1ea43.

[7] Mats J. Stensrud and Miguel A. Hernán. Why Test for Proportional Hazards? JAMA. 2020; 323 (14):1401–1402, 04. ISSN 0098-7484. doi: 10.1001/jama.2020.1267. URL https://doi.org/10.1001/jama.2020.1267.

[8] Daniel Westreich and Sander Greenland. The Table 2 Fallacy: Presenting and Interpreting Confounder and Modifier Coefficients. American Journal of Epidemiology. 2013;177(4):292–298, 01. ISSN 0002-9262. doi: 10.1093/aje/kws412. URL https://doi.org/10.1093/aje/kws412.

[9] Gary Smith. Step away from stepwise. Journal of Big Data. 2018; 5(1):1–12. https://doi.org/10.1186/s40537-018-0143-6

[10] Miguel A. Hernán and James M. Robins. Using Big Data to Emulate a Target Trial When a Randomized Trial Is Not Available. American Journal of Epidemiology. 2016;183(8):758–764, 03. ISSN 0002-9262. doi: 10.1093/aje/kwv254. URL https://doi.org/10.1093/aje/kwv254.

[11] Maya L. Petersen and Mark J. van der Laan. Causal models and learning from data: Integrating causal modeling and statistical estimation. Epidemiology. 2014;25(3):418–426. ISSN 10443983. URL http://www.jstor.org/stable/24759134.

[12] Jeremy A Labrecque and Sonja A Swanson. Target trial emulation: teaching epidemiology and beyond. European journal of epidemiology. 2017;32(6):473–475.

[13] Miguel A. Hernán Brian C. Sauer, Sonia Hernández-Díaz, Robert Platt, and Ian Shrier. Specifying a target trial prevents immortal time bias and other self-inflicted injuries in observational analyses. Journal of Clinical Epidemiology. 2016;79:70–75. ISSN 0895-4356. doi: https://doi.org/10.1016/j.jclinepi.2016.04.014. URL https://www.sciencedirect.com/science/article/pii/S0895435616301366.

[14] Jonathan AC Sterne, Srinivas Murthy, Janet V Diaz, Arthur S Slutsky, Jesús Villar, Derek C Angus, et al. Association between administration of systemic corticosteroids and mortality among critically ill patients with covid-19: a meta-analysis. Jama.. 2020;324(13):1330–1341.

[15] von Elm E, Altman DG, Egger M, Pocock SJ, Gøtzsche PC, Vandenbroucke JP; STROBE Initiative. The Strengthening the Reporting of Observational Studies in Epidemiology (STROBE)statement: guidelines for reporting observational studies. J Clin Epidemiol. 2008 Apr;61(4):344–9. PMID: 18313558

[16] Steroid conversion calculator. URL https://www.mdcalc.com/steroid-conversion-calculator. Accessed May 10, 2020.

[17] Parag Goyal, Justin J Choi, Laura C Pinheiro, Edward J Schenck, Ruijun Chen, Assem Jabri, et al. Clinical characteristics of covid-19 in new york city. New England Journal of Medicine. 2020;382(24): 2372–2374.

[18] Edward J Schenck, Katherine L Hoffman, Marika Cusick, Joseph Kabariti, Evan T Sholle, and Thomas R Campion Jr. Critical care database for advanced research (cedar): An automated method to support intensive care units with electronic health record data. Journal of Biomedical Informatics. 2021;118:103789.

[19] Bibhas Chakraborty and EE Moodie. Statistical methods for dynamic treatment regimes. Springer-Verlag. 2013. doi, 10:978–1.

[20] James Robins. A new approach to causal inference in mortality studies with a sustained exposure period—application to control of the healthy worker survivor effect. Mathematical Modelling. 1986;7(9):1393–1512. ISSN 0270-0255. doi: https://doi.org/10.1016/0270-0255(86)90088-6. URL https://www.sciencedirect.com/science/article/pii/0270025586900886.

[21] Alexander R Luedtke, Oleg Sofrygin, Mark J van der Laan, and Marco Carone. Sequential double robustness in right-censored longitudinal models. arXiv preprint arXiv. 2017;1705.02459.

[22] Iván Díaz, Nicholas Williams, Katherine L. Hoffman, and Edward J. Schenck. Nonparametric causal effects based on longitudinal modified treatment policies. Journal of the American Statistical Association. 2021;0(0):1–16. doi: 10.1080/01621459.2021.1955691. URL https://doi.org/10.1080/01621459.2021.1955691.

[23] Mark J. van der Laan and Sherri Rose. Targeted Learning in Data Science: Causal Inference for Complex Longitudinal Studies. Springer Publishing Company, Incorporated, 1st edition, 2018. ISBN 3319653032.

[24] Victor Chernozhukov, Denis Chetverikov, Mert Demirer, Esther Duflo, Christian Hansen, Whitney Newey, et al. Double/debiased machine learning for treatment and structural parameters. The Econometrics Journal. 2018;21(1):C1–C68, 01. ISSN 1368-4221. doi: 10.1111/ectj.12097. URL https://doi.org/10.1111/ectj.12097.

[25] T. A. Gerds and M. Schumacher. On functional misspecification of covariates in the Cox regression model. Biometrika. 2001;88(2):572–580, 06. ISSN 0006-3444. doi: 10.1093/biomet/88.2.572. URL https://doi.org/10.1093/biomet/88.2.572.

[26] Leo Breiman. Stacked regressions. Machine learning. 1996;24(1):49–64.

[27] Mark J. van der Laan, Eric C Polley, and Alan E. Hubbard. Super learner. Statistical Applications in Genetics and Molecular Biology. 2007;6(1). doi: doi:10.2202/1544-6115.1309. URL https://doi.org/10.2202/1544-6115.1309.

[28] Faegheh Ebrahimi Chaharom, Leili Pourafkari, Ali Asghar Ebrahimi Chaharom, and Nader D Nader. Effects of corticosteroids on covid-19 patients: A systematic review and meta-analysis on clinical outcomes. Pulmonary pharmacology & therapeutics, 2021;page 102–107.

[29] Bruno Martins Tomazini, Israel Silva Maia, Flavia Regina Bueno, Maria Vitoria Aparecida Oliveira Silva, Franca Pellison Baldassare, et al. Covid-19-associated ards treated with dexamethasone (codex): study design and rationale for a randomized trial. Revista Brasileira de terapia intensive. 2020;32:354–362.

[30] Maryam Edalatifard, Maryam Akhtari, Mohammadreza Salehi, Zohre Naderi, Ahmadreza Jamshidi, Shayan Mostafaei, et al. Intravenous methylprednisolone pulse as a treatment for hospitalised severe covid-19 patients: results from a randomised controlled clinical trial. European Respiratory Journal. 2020;56(6).

[31] Luis Corral-Gudino, Alberto Bahamonde, Francisco Arnaiz-Revillas, Julia Gómez-Barquero, Jesica Abadía-Otero, Carmen García-Ibarbia, et al. Methylprednisolone in adults hospitalized with covid-19 pneumonia. Wiener klinische Wochenschrift. 2021;133(7):303–311.

[32] Carina Wagner, Mirko Griesel, Agata Mikolajewska, Anika Mueller, Monika Nothacker, Karoline Kley, et al. Systemic corticosteroids for the treatment of covid-19. Cochrane Database of Systematic Reviews. 2021;(8).

[33] Ana Fernández-Cruz, Belén Ruiz-Antorán, Ana Muñoz-Gómez, Aránzazu Sancho-López, Patricia Mills-Sánchez, Gustavo Adolfo Centeno-Soto, et al. A retrospective con-’ s trolled cohort study of the impact of glucocorticoid treatment in sars-cov-2 infection mortality. Antimicrobial agents and chemotherapy, 2020;64(9):e01168–20.

[34] Brian C Nelson, Justin Laracy, Sherif Shoucri, Donald Dietz, Jason Zucker, Nina Patel, et al. Clinical outcomes associated with methylprednisolone in mechanically ventilated patients with covid-19. Clinical Infectious Diseases. 2021;72(9):e367–e372.

[35] Jiao Liu, Sheng Zhang, Xuan Dong, Zhongyi Li, Qianghong Xu, Huibin Feng, et al. Corticosteroid treatment in severe covid-19 patients with acute respiratory distress syndrome. The Journal of clinical investigation. 2020;130(12):6417–6428.

[36] Linda E Levesque, James A Hanley, Abbas Kezouh, and Samy Suissa. Problem of immortal time bias in cohort studies: example using statins for preventing progression of diabetes. Bmj. 2010;340.

[37] Michele Bartoletti, Lorenzo Marconi, Luigia Scudeller, Livia Pancaldi, Sara Tedeschi, Maddalena Giannella, et al. Efficacy of corticosteroid treatment for hospitalized patients with severe covid-19: a multicentre study. Clinical Microbiology and Infection. 2021;27(1):105–111.

[38] Jesús Rodríguez-Baño, Jerónimo Pachón, Jordi Carratalá, Pablo Ryan, Inmaculada Jarrín, María Yllescas, et al. Treatment with tocilizumab or corticosteroids for covid-19 patients with hyperinflammatory state: a multicentre cohort study (sam-covid-19). Clinical Microbiology and Infectio. 2021;27(2):244–252.

[39] Ivan Cusacovich, Alvaro Aparisi, Miguel Marcos, Cristina Ybarra-Falcon, Carolina Iglesias-Echevarria, et al. Corticosteroid pulses for hospitalized patients with covid-19: effects on mortality. Mediators of inflammation, 2021.

[40] Cecilia Tortajada, Enrique Colomer, Juan C Andreu-Ballester, Ana Esparcia, Carmina Oltra, and Juan Flores. Corticosteroids for covid-19 patients requiring oxygen support? yes, but not for everyone: effect of corticosteroids on mortality and intensive care unit admission in patients with covid-19 according to patients’ oxygen requirements. Journal of Medical Virology. 2021;93(3):1817–1823.

[41] Gareth J Griffith, Tim T Morris, Matthew J Tudball, Annie Herbert, Giulia Mancano, Lindsey Pike, et al. Collider bias undermines our understanding of covid-19 disease risk and severity. Nature communications. 2020;11(1):1–12.

