## Appendix for "Comparison of a Target Trial Emulation Framework to Cox Regression to Estimate the Effect of Corticosteroids on COVID-19 Mortality"

**Table of Contents:**

**eMethods**

1 Intention-to-treat versus per-protocol effect
2 Choice of study time frame
3 Confounders
4 Decision to bin data in 24-hour windows
5 Informative measurement
6 Estimand
7 Transportability
8 Data analysis plan
9 Analytical file
10 Software

**eResults**

1 Participants
2 Informative measurement
3 Sensitivity analysis: Removing co-treatments as confounders
4 Sensitivity analysis: MICE

**eTables**

eTable 1. RECOVERY study design comparison

eTable 2. RECOVERY study population comparison

eTable 3. RCT meta-analysis corticosteroid intervention comparison

eTable 4. Corticosteroid types, co-treatments, and discharge pathways.

eTable 5. Informative measurement over time

**eFigures**

eFigure 1. Treatment timelines for 50 patients

eFigure 2. Illustration of analytical data structure

**eMethods**

1 **Per-protocol effect versus intention-to-treat**

Intention-to-treat, or the comparison of patients who were randomized to each treatment arm regardless of whether they actually received the assigned treatment, is the primary estimand of interest in many RCTs. However, it is rarely possible to emulate this contrast using observational data.^1^ The closest possible analog of an intention-to-treat design for our corticosteroid question may have been analyzing the difference in patients who were prescribed corticosteroids, whether or not they were actually administered, if that data were available.^1^

Instead of pursuing intention-to-treat, we design a hypothetical target trial primarily interested in the per-protocol effect, or the difference in patients who actually follow each of the regimes of interest. To analyze the closest possible analog of a per-protocol effect, we will account for baseline and post-baseline (time-dependent) confounders in the administration of corticosteroid treatment over time, since corticosteroid administration and adherence depends on changing physiological characteristics of patients.^1^ See “Using Big Data to Emulate a Target Trial When a Randomized Trial Is Not Available” by Hernan et al. for a more detailed description of the intention-to-treat versus per-protocol comparisons in target trials.^1^

2 **Study time frame**

The time frame and location of this target trial emulation, March-May 2020 at a large hospital system in New York City, lends itself well to studying the effect of corticosteroids on mortality. The dates of the study are during the peak of NYC’s initial pandemic surge conditions, meaning there were thousands of patients admitted within a short period of time, all infected with the same COVID-19 variant and treated with a relatively equal, albeit limited, level of provider knowledge on the disease. This time period was prior to vaccines and other treatments now known to be effective, such as monoclonal antibodies or antiviral treatments. A key aspect of this time period is that the effectiveness of corticosteroids was not yet established; in fact, there was some evidence early in the pandemic that corticosteroids could be harmful. The largest and most influential study, Randomized Evaluation of COVID-19 Therapy (RECOVERY), published preliminary results in June 2020^2^ and significantly changed clinical practice afterwards. Our pre-RECOVERY time period resulted in high variability in provider practice regarding the timing and administration of steroids, as shown in eFigure 1. Although variability in provider practice can complicate simpler analyses of observational data, it is ideal for estimating causal effects in combination with an advanced study design because there is natural experimentation already occurring in the data.

3 **Confounders**

The full list of baseline confounders includes age, sex, race, ethnicity, body mass index (BMI), comorbidities (cerebral vascular event, hypertension, diabetes mellitus, cirrhosis, chronic obstructive pulmonary disease, active cancer, asthma, interstitial lung disease, chronic kidney disease, immunosuppression, human immunodeficiency virus (HIV)-infection, and home oxygen use), hospital admission location, and mode of respiratory support within 3 hours of hospital admission. Time-dependent confounders include vital signs (heart rate, pulse oximetry percentage, respiratory rate, temperature, systolic/diastolic blood pressure), laboratory results (blood urea nitrogen (BUN)-creatinine ratio, creatinine, neutrophils, lymphocytes, platelets, bilirubin, blood glucose, D-dimers, C-reactive protein, activated partial thromboplastin time, prothrombin time, arterial partial pressures of oxygen and carbon dioxide), supplemental oxygen, and concurrent pharmaceutical treatments. Supplemental oxygen support was defined by no supplemental oxygen, non-invasive mechanical ventilation (IMV) supplemental oxygen, and IMV.

The concurrent treatments included as confounders were vasopressors, diuretics, Angiotensin-converting enzyme (ACE) and Angiotensin receptor blockers (ARBs), hydroxychloroquine, and tocilizumab. Of note, none of these treatments have been established as effective therapies for COVID-19. In addition, physiological changes from these treatments should be captured in the plethora of vital signs and laboratory results we include as time-varying confounders. A sensitivity analysis without these experimental treatments as confounders is provided in the eResults, section 3.

4 **Decision to bin data in 24-hour time intervals**

This study required binning our data into time intervals, since we needed a standard time to recalibrate dosing corticosteroid dosing information across patients. For this research question, we chose to bin the data in 24-hour intervals. In cases of multiple laboratory results or vital signs in a 24-hour period, the clinically worst value was used.

The 24-hour window was specified *a priori* by clinical guidance: we defined a corticosteroid exposure as a methylprednisolone-equivalent reaching >0.5 mg/kg body weight over a 24-hour period. Thus, patients could “qualify” as having a corticosteroids exposure up until the end of each 24-hour binning period. The 24-hour time period is how all of the RCTs used in the WHO meta-analysis also define their treatment regimes.

The biological reason we (and also presumably the RCTs) used for choosing 24 hours is that it should adequately capture the mechanistic action timeline of corticosteroids. In a randomized controlled trial (RCT) looking at hydrocortisone for mechanically ventilated septic shock patients, the mean arterial pressure between hydrocortisone and placebo patients does not begin to noticeably diverge until study day 2 (Figure S4b of Adrenal et al.).^3^ This led us to the decision that 24-hour windows were sufficient to capture the physiological dynamics for corticosteroids.

We can definitively say that the outcome of death occurred after the covariates and exposures were measured on a causal pathway. However, this binning strategy of covariates and exposures could theoretically lead to measured time-varying covariates (laboratory tests or vital signs) occurring concurrently or posteriorly to corticosteroid administration. The impact on this analysis of the data should be minimal, especially due to the relatively slower biological time of action time of corticosteroids.

5 **Informative measurement**

The measurement, or lack thereof, of covariates in this analysis is often highly prognostic. The analytical strategy for handling missing variables was to fully account for the measurement process by including an indicator of whether the variable was measured. Missing baseline categorical variables were handled using an indicator for informative measurement (e.g., missing race was its own category). Missing time-varying continuous variables were also handled with an indicator for informative measurement. In order to make the software work for estimating our dynamic regimes, we then substituted unmeasured continuous variables with an arbitrary value (median). Of note, the same results could be obtained with any arbitrary value, such as 9999, after adding the indicator of the measurement process.

Further details on the informative measurement process are shown in eTable 5. A sensitivity analysis in which Multiple Imputation Chained Equations^4^ (MICE) was used sequentially at every time point to substitute missing values (after including the informative measurement variable) was also performed. These results are in eResults, section 4.

6 **Estimand**

The g-computation formula can be conceptualized as a sequential regression of the outcome as follows.^5^ First, a model for the probability of mortality a day 28 is estimated amongst patients who have not been lost to follow-up, with predictors given by all the data prior to day 28. This model fit is then used to produce predictions of the probability of mortality for all patients had the hypothetical intervention of interest been implemented at day 28. This probability is then used as a pseudo-outcome in a regression model where the predictors are all variables measured up to day 27. This model is then used to obtain new predictions for all patients had the hypothetical intervention of interest been implemented at day 27. This process is iterated until day 1, at which point the predictions are averaged. This average is the estimated mortality rate under the hypothetical treatment rule.

The G-computation formula is one possible way to identify our estimand of interest. A key assumption when estimating the g-computation formula is that treatment and loss-to-follow-up are independent of the counterfactual outcome at every time point conditional on measured confounders. Other identification strategies which do not require unconfoundedness, but which we did not use in this application, include double negative controls, identification of bounds, sensitivity analyses, or front-door criterion.^6-9^

7 **Transportability**

The comparability of our hypothetical target trial to the RCTs in the WHO working group’s corticosteroid meta-analysis as a benchmark is not exact.^10^ This is partially because we did not have the sample size to look at individual corticosteroid types, such as dexamethasone or hydrocortisone. It is unknown how the heterogeneity in treatment regimes between the trials and our cohort may affect our use of the WHO’s meta-analysis as a benchmark. We summarize the treatment regimes for the seven RCT’s included in the WHO meta-analysis in eTable 1.

It is also unclear how differences in patient eligibility and participation characteristics may affect the use of the meta-analysis as a benchmark. We show characteristics of RECOVERY’s study design and patient characteristics (eTable 2 and 3) as a reference to demonstrate how the RCTs compare to our own research. Although our hypothetical intervention did generally resemble the RCTs in overall study design, we looked at different contrasts of interest (see Section 1 – Intention to treat versus per-protocol effect). There are also some differences in study population, notably race and some comorbidities. Differences such as study location (e.g. RECOVERY took place in the United Kingdom) may also play a role in comparability of our results from a study cohort in New York City, either from RCTs to the general population of COVID-19 patients, or between our study and the RCTs.

Our hypothetical target trial protocol also deviates, as discussed in Section 1 of the eMethods (Intention-to-treat versus per-protocol estimand), from the RCTs because it does not allow for physicians to discontinue corticosteroids before the assigned six days, or for patients in the “standard of care” arm to receive corticosteroids as part of standard of care. We are measuring the effect of patients receiving and continue to receive their assigned treatment. This may also limit comparability, since the RCTs were often interested in intention-to-treat as the primary outcome. However, patients in the RCTs who were given corticosteroids when assigned to the treatment arm or not given corticosteroids when assigned should bias results towards the null. These comparability issues, which are discussed under the names of transportability, generalizability, and internal/external validity in the causal inference literature, are open problems and require analytical tools outside the scope of this paper.^11^

8 **Data analysis plan**

The Sequentially Double Robust (SDR) estimator uses the probability of receiving the intervention, the probability of experiencing the outcome, and the probability of censoring at each time point to estimate the final survival probability under a given intervention. For longitudinal studies, the SDR estimator estimates these probabilities iteratively at discrete time points. At each time point, only the intervention or outcome model need be correctly specified for the final estimate to be consistent.^12^ We fit each model using an ensemble machine learning via the super learner algorithm.^13,14^ The super learner algorithm creates a weighted combination of algorithms in a user-supplied library to obtain a final prediction that is guaranteed to outperform each algorithm in the library as sample size grows.^14^

We use a super learner library containing candidate learners of multivariate adaptive regression splines (MARS) with varying parameters, Bayesian Additive Regression Trees (BART), penalized linear regression with penalties of 0, 0.5, and 1 (Ridge, Elastic Net, and LASSO), and an intercept only (mean) model. To fit these regressions, we assume that outcomes at day *t* only depend on baseline confounders and time-varying covariates measured in the previous two days. We employ 10-fold cross-validation on the super learner estimation and 10-fold cross-fitting on the SDR estimator.

Hypothesis tests for both the target trial emulation and model-first approaches are two-sided with a Type I error rate.

9 **Analytical file**

eFigure 2 shows an illustrated example of the analytical file. Time-dependent confounders, censoring indicators, and outcome indicators are binned in 24-hour windows from time of randomization. The final analytical file contains one row per patient with one column for each baseline confounder and one column for each time-dependent confounder, outcome, and censoring indicator at all 28 time points. Further details on the analytical file creation are publicly available in a working Github tutorial: <https://github.com/kathoffman/steroids-trial-emulation>. A template for creating the illustrative figures from this report is also included on the Github repository.

10 **Software**

Data were analyzed in R version 4.0.3 with the open-source packages ggplot2, gtsummary, survival, survminer, mice, sl3, and lmtp.^15-22^

**eResults**

1 **Participants**

There were 3,737 unique adult COVID-19 patients admitted during the study time frame. In cases of multiple patient-encounters, the first visit was used. 165 patients were excluded due to transfer in from an outside hospital. An additional 274 were excluded due to chronic corticosteroid use, leaving a final cohort of 3,298.

2 **Informative measurement**

Of the baseline confounders, Body Mass Index (BMI) was missing for 5.8% patients, race was missing for 218 (6.6%), and ethnicity was missing for 596 (18%). All other baseline variables were complete. For the time-varying covariates, vital signs, co-treatment, and supplemental oxygen data were complete. For routine laboratory results such as platelets, nearly all patients had complete data (e.g. 90% completeness for day 0 platelets). Measurement of other COVID-specific laboratory results, e.g. D-dimers, was less common and highly prognostic as discussed in eMethods section 5, Informative measurement. A table showing the percentage of the at-risk population with informative measurement for the time-varying covariates is provided in eTable 5.

3 **Sensitivity Analysis 1: Removing co-treatments as confounders**

A sensitivity analysis removing co-treatments as confounders yielded the result that under the no corticosteroids regime, 28-day mortality is estimated to be 32.2% (30.8-33.5). Under the corticosteroids regime, 28-day mortality is estimated to be 25.9% (24.7-27.0%). This yields an estimated decrease in 28-day mortality of 6.3% (5.4-7.2).

4 **Sensitivity Analysis 2: MICE**

A sensitivity analysis using MICE to sequentially substitute missing values at every time point after adding a variable for informative missingness yielded the result that under the no corticosteroids regime, 28-day mortality is estimated to be 32.9% (31.7-34.2). Under the corticosteroids regime, 28-day mortality is estimated to be 26.4% (25.4-27.5). This yields an estimated decrease in 28-day mortality of 6.5% (5.7-7.3).

**eTables**

**eTable 1.** A comparison of study designs between the largest RCT in the WHO corticosteroid meta-analysis, the Randomized Evaluation of Covid-19 Therapy (RECOVERY) dexamethasone trial, and the hypothetical target trial’s emulation.

|  | **RECOVERY dexamethasone trial^a^** | **Hypothetical target trial emulation** |
| --- | --- | --- |
| **Eligibility criteria** | Adult hospitalized COVID-19 patients without a medical history that may, in the opinion of the attending clinician, put the patient at substantial risk if they were to participate in the trial | Adult hospitalized COVID-19 patients not receiving corticosteroids for chronic use or transferring in from an outside hospital |
| **Treatment arms** | Low-dose corticosteroids (dexamethasone 6 mg) once daily for 10 days at physician’s discretion plus standard of care vs. standard of care without corticosteroids | Corticosteroids >0.5 mg/kg body weight per day for 6 days from time of severe hypoxia plus standard of care vs. standard of care without corticosteroids |
| **Assignment strategies** | Subjects randomly assigned upon enrollment (which may occur days after hospitalization) | Subjects randomly assigned upon enrollment (hospitalization admission) |
| **Follow-up period** | Begins at randomization and ends at 28 days, death, or loss to follow up, whichever comes first | Begins at randomization and ends at 28 days, death, or loss to follow up, whichever comes first |
| **Outcome** | 28-day mortality | 28-day mortality |
| **Causal contrasts of interest** | Intention-to-treat | Per-protocol |
| ^a^We present RECOVERY as the comparative corticosteroid RCT as it was the largest and most influential to the WHO working group’s meta-analysis results. ^b^After May 9, 2020, the adult age restriction was removed. | | |

**eTable 2.** RECOVERY study population comparison

| **Characteristic** | **RECOVERY dexamethasone trial^a^** | **Hypothetical target trial emulation** |
| --- | --- | --- |
| **Age^b^** | 66.9 (15) | 64.2 (17) |
| **Sex** |  |  |
| **Male** | 64% | 60% |
| **Female** | 36% | 40% |
| **Race** |  |  |
| **White** | 74% | 28% |
| **Non-white or unknown** | 26% | 72% |
| **Diabetes** | 25% | 31% |
| **Human Immunodeficiency Virus** | 1% | 1% |
| **Heart disease** | 28% | 14% |
| ^a^We present RECOVERY as the comparative corticosteroid RCT as it was the largest and most influential to the WHO working group’s meta-analysis results. ^b^To match the RECOVERY table, mean (standard deviation) is shown for continuous variables. | | |

**eTable 3.** Summary of corticosteroid interventions of the seven RCTs included in the WHO working group’s meta-analysis.

| **Trial (ClinicalTrials.gov Identifier)** | **Corticosteroids Treatment Regimen** |
| --- | --- |
| RECOVERY (NCT04381936) | Dexamethasone 6 mg once daily for 10 days at physician’s discretion plus standard of care vs. standard of care without corticosteroids |
| CoDEX (NCT04327401) | Dexamethasone 20mg intravenous (IV) daily for 5 days, followed by 10mg IV daily for 5 days |
| DEXA‐COVID19 (NCT04325061) | 20 mg dexamethasone IV daily from Day 1 of randomization during 5 days, followed by 10 mg IV daily from Day 6 to 10 of randomization |
| CAPE_COVID (NCT02517489) | Hydrocortisone given in a double-blind fashion for 8 or 14 full days via IV. The treatment course will include 4 or 7 days of full dose (200 mg/day by continuous infusion), 2 or 4 days of half dose (100 mg/day by continuous infusion), and 2 or 3 days of tapering dose (50 mg/day by continuous infusion). Duration of treatment is chosen upon patient initial improvement. |
| COVID STEROID (NCT04348305) | Hydrocortisone given via continuous infusion (200 mg (104 ml) every 24 hours) or Bolus injections (50 mg (10 ml) every 6 hours) for 7 days |
| REMAP‐CAP (NCT02735707) | 50 or 100mg hydrocortisone administered IV every 6 hours for up to 7 days |
| Steroids‐SARI (NCT04244591) | Methylprednisolone 40 mg every 12 hours for 5 days |

**eTable 4.** Co-treatments and corticosteroid types received ever during study follow up, and loss to follow-up pathways.

| **Characteristic^a^** | **N=3,298** |
| --- | --- |
| **Corticosteroid type^b^** |  |
| **Dexamethasone** | 74 (2.2%) |
| **Hydrocortisone** | 100 (3%) |
| **Methylprednisolone** | 306 (9.3%) |
| **Prednisolone** | 2 (0.1%) |
| **Prednisone** | 67 (2%) |
| **Co-treatments^c^** |  |
| **ACE/ARBs** | 263 (8.0%) |
| **Diuretics** | 501 (15%) |
| **Inotropes** | 42 (1.3%) |
| **Interferon beta-1a** | 0 (0%) |
| **Intravenous immunoglobulin therapy** | 10 (0.3%) |
| **Plaquenil** | 2,265 (69%) |
| **Sarilumab** | 16 (0.5%) |
| **Tamiflu** | 70 (2.1%) |
| **Tocilizumab** | 131 (4.0%) |
| **Vasopressors** | 538 (16%) |
| **Loss the follow-up pathway** |  |
| **Acute Rehab** | 39 (1.7%) |
| **Home** | 1,731 (76%) |
| **Hospice** | 24 (1.1%) |
| **Other^d^** | 51 (2.2%) |
| **Shelter** | 16 (0.7%) |
| **Skilled Nursing Facility** | 261 (11%) |
| **Subacute Rehab** | 104 (4.6%) |
| ^a^Abreviations: ACE/ARBs = Angiotensin-converting enzyme / Angiotensin receptor blockers. ^b^Corticosteroid types are not mutually exclusive. Only exposures greater than 0.5 mg/kg body weight per 24 hours are shown. ^c^Co-treatments refer to the patient receiving more than two consecutive days of the co-treatments at any point during study follow up. Experimental treatments in place during the study time are shown, despite some patients not receiving any. ^d^Other includes hotel, group home, undomiciled, psychiatric facility, or left against medical advice. | |

**eTable 5.** Informative measurement percentages by study day. Percentage refers to the proportion of patients with informative measurement in the at-risk group.

| **Study Day** | **Informative Measurement (%)** |
| --- | --- |
| **0** | 73.3 |
| **1** | 79.7 |
| **2** | 77.9 |
| **3** | 77.3 |
| **4** | 74 |
| **5** | 73 |
| **6** | 71 |
| **7** | 68.9 |
| **8** | 66.4 |
| **9** | 65.9 |
| **10** | 66.4 |
| **11** | 66.3 |
| **12** | 66.6 |
| **13** | 63.2 |
| **14** | 62.7 |
| **15** | 61.4 |
| **16** | 65.9 |
| **17** | 64.8 |
| **18** | 62.5 |
| **19** | 64.1 |
| **20** | 66.1 |
| **21** | 62.6 |
| **22** | 66.9 |
| **23** | 65.4 |
| **24** | 61.6 |
| **25** | 67.4 |
| **26** | 65.9 |
| **27** | 63.3 |

**eFigures**

**eFigure 1.** Treatment timelines for 50 patients. A random sample of 50 patients’ observed hospital courses, ordered by hospital length of stay. Each line represents one patient, and the color of the line represents that patient’s intubation status (a time-dependent confounder to corticosteroids administration and death). A red circle indicates the time of severe hypoxia. Yellow squares indicate steroids administration. Patients received corticosteroids at various times and durations throughout their hospitalization course and in relation to meeting the criteria of severe hypoxia, which is ideal for meeting natural experimentation conditions.

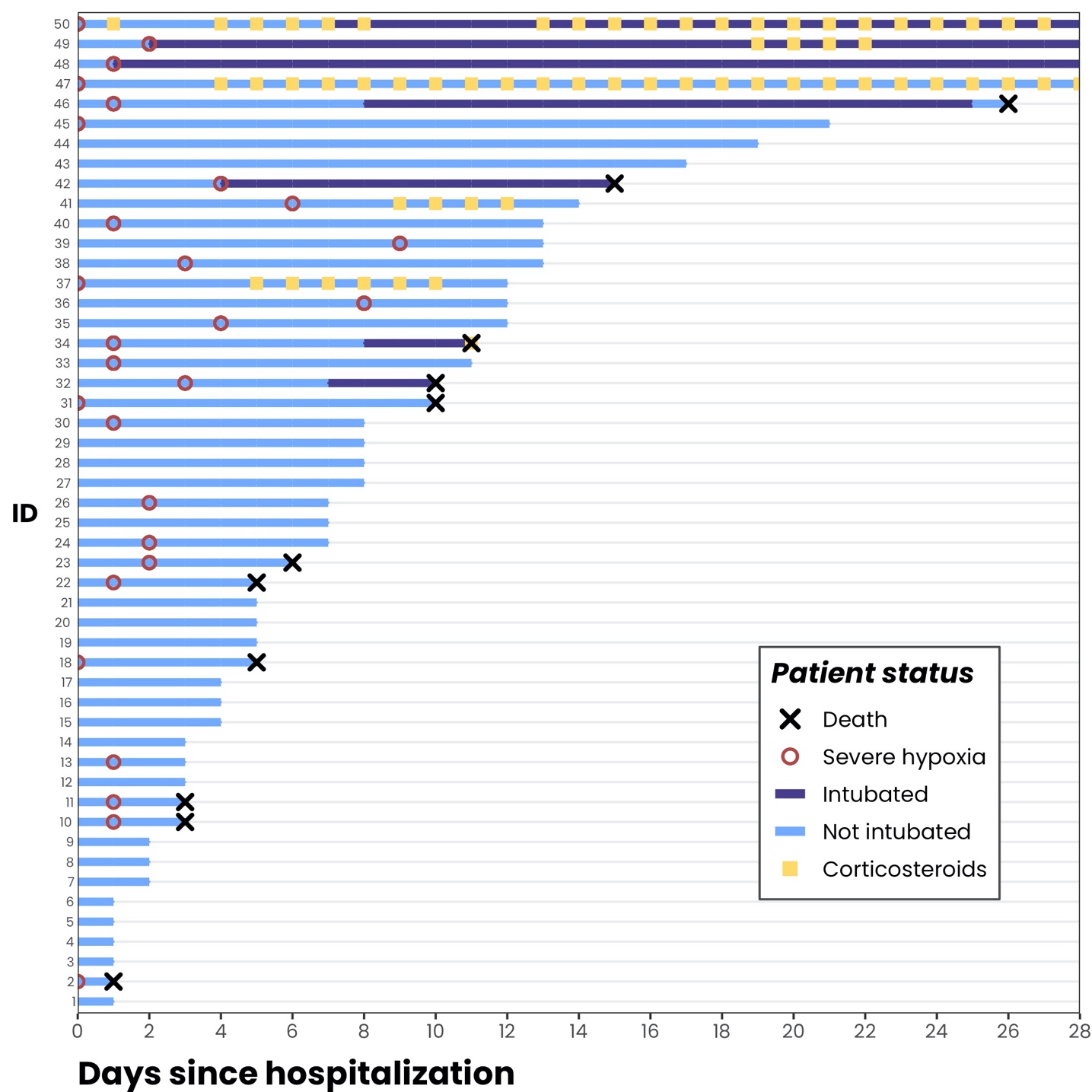

**eFigure 2.** Illustration of the analytical data file. The analytical file contained one row per patient and one column per baseline confounder. For every time point, there was one column per time-dependent confounder, a treatment status indicator, censoring indicator, and outcome indicator. The data frame was prepared in R and used in combination with the open-source R package lmtp.

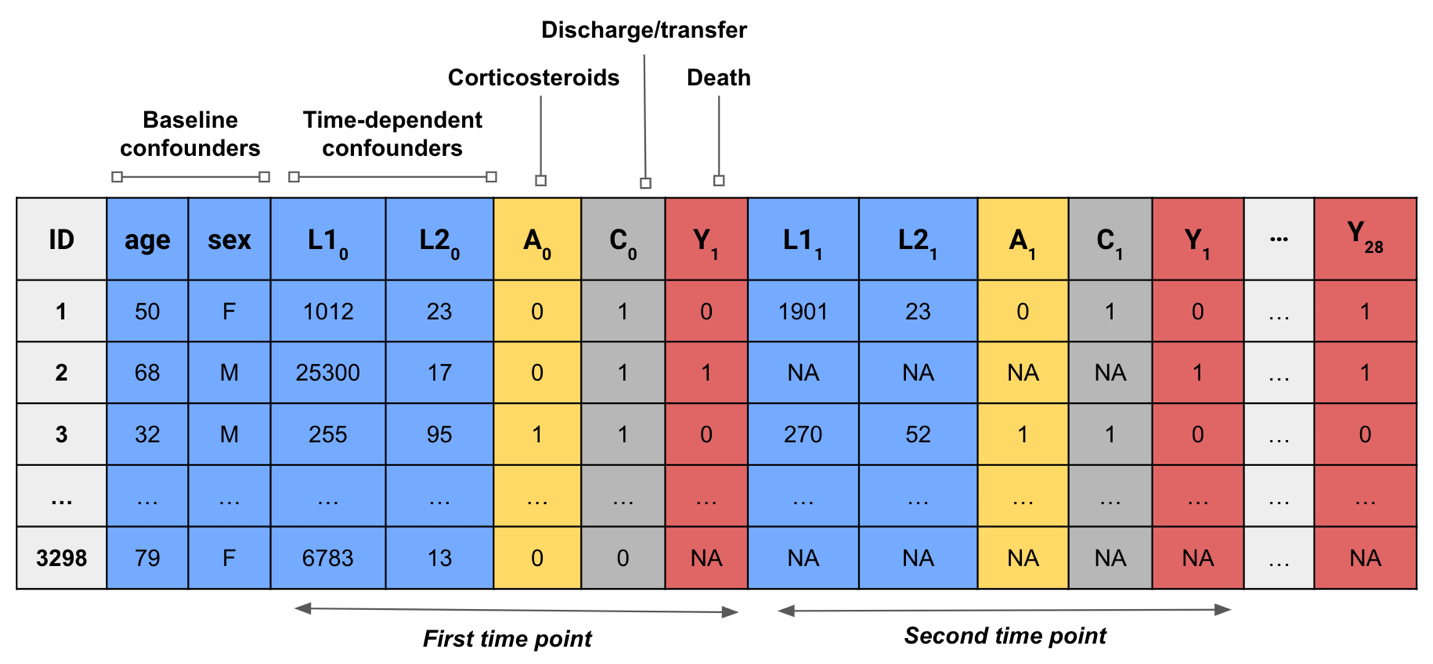

**References**

[1] Miguel A. Hernán and James M. Robins. Using Big Data to Emulate a Target Trial When a Randomized Trial Is Not Available. *American Journal of Epidemiology*. 2016;183(8):758–764. ISSN 0002-9262. doi: 10.1093/aje/kwv254. DOI: [https://doi.org/10.1093/ aje/kwv254.](https://doi.org/10.1093/aje/kwv254)
[2] Faust, S., Horby, P., Lim, W. S., Emberson, J., Mafham, M., Bell, J. L., et al. Effect of dexamethasone in hospitalized patients with COVID-19: preliminary report. *MedRxiv*. 2020. DOI: 10.1056/NEJMoa2021436
[3] Venkatesh, B., Finfer, S., Cohen, J., Rajbhandari, D., Arabi, Y., Bellomo, R., Billot, L., Correa, M., Glass, P., Harward, M. and Joyce, C., 2018. Adjunctive glucocorticoid therapy in patients with septic shock. *New England Journal of Medicine.* 2018;378(9), pp.797-808. DOI: 10.1056/NEJMoa1705835
[4] Heejung Bang and James M Robins. Doubly robust estimation in missing data and causal inference models. *Biometrics.* 61(4):962–973.
[5] Robins JM, Hernán MA. *Causal Inference: What If*. Boca Raton: Chapman & Hall, 2020.
[6] Rosenbaum PR. The role of known effects in observational studies. *Biometrics*. 1989;45, 557–569.
[7] Jo B, Vinokur AD. Sensitivity Analysis and Bounding of Causal Effects With Alternative Identifying Assumptions. *J Educ Behav Stat.* 2011 Aug;36(1):415-440. doi: 10.3102/1076998610383985. PMID: 21822369; PMCID: PMC3150587.
[8] Vanderweele TJ, Arah OA. Bias formulas for sensitivity analysis of unmeasured confounding for general outcomes, treatments, and confounders. *Epidemiology.* 2011 Jan;22;(1):42-52. doi: 10.1097/EDE.0b013e3181f74493. PMID: 21052008; PMCID: PMC3073860.
[9] Fulcher, Isabel R., et al. "Robust inference on population indirect causal effects: the generalized front door criterion." *Journal of the Royal Statistical Society: Series B (Statistical Methodology)*  2020; 82(1):199-214.
[10] Dahabreh IJ, Robins JM, Hernán MA. Benchmarking Observational Methods by Comparing Randomized Trials and Their Emulations. *Epidemiology.* 2020; Sep;31(5):614-619. doi: 10.1097/EDE.0000000000001231. PMID: 32740470.
[11] Degtiar, Irina, and Sherri Rose. "A review of generalizability and transportability." *arXiv preprint arXiv.* 2021;*2102.11904*.
[12] Iván Díaz, Nicholas Williams, Katherine L. Hoffman, and Edward J. Schenck. Nonparametric causal effects based on longitudinal modified treatment policies. *Journal of the American Statistical Association*. 2021;0(0):1–16. doi: 10.1080/01621459.2021.1955691. URL [https://doi.org/10.1080/01621459.2021.1955691.](https://doi.org/10.1080/01621459.2021.1955691)
[13] Leo Breiman. Stacked regressions. *Machine learning*. 1996;24(1):49–64.
[14] Mark J. van der Laan, Eric C Polley, and Alan E. Hubbard. Super learner. *Statistical Applications in Genetics and Molecular Biology*. 2007;6(1). doi: doi:10.2202/1544-6115.1309. URL [https://doi.org/10.2202/1544-6115.1309.](https://doi.org/10.2202/1544-6115.1309)
[15] R Core Team (2022). R: A language and environment for statistical computing. R Foundation for Statistical Computing, Vienna, Austria.URL https://www.R-project.org/.
[16] Hadley Wickham. *ggplot2: Elegant Graphics for Data Analysis*. Springer-Verlag New York, 2016. ISBN 978-3-319-24277-4. URL [https://ggplot2.tidyverse.org.](https://ggplot2.tidyverse.org/)
[17] Daniel D. Sjoberg, Karissa Whiting, Michael Curry, Jessica A. Lavery, and Joseph Larmarange. Reproducible summary tables with the gtsummary package. *The R Journal*, 13:570–580, 2021. doi: 10.32614/RJ-2021-053. URL [https://doi.org/10.32614/ RJ-2021-053.](https://doi.org/10.32614/RJ-2021-053)
[18] Terry M Therneau. *A Package for Survival Analysis in R*, 2021. URL [https://CRAN. R-project.org/package=survival.](https://CRAN.R-project.org/package=survival) R package version 3.2-13.
[19] Alboukadel Kassambara, Marcin Kosinski, and Przemyslaw Biecek. *survminer: Drawing*

*Survival Curves using ’ggplot2’*, 2021. URL [https://CRAN.R-project.org/package= survminer.](https://CRAN.R-project.org/package=survminer) R package version 0.4.9.

[20] van Buuren S, Groothuis-Oudshoorn K (2011). “mice: Multivariate Imputation by Chained Equations in R.” *Journal of Statistical Software*, **45**(3), 1-67. doi: [10.18637/jss.v045.i03](https://doi.org/10.18637/jss.v045.i03).
[21] Jeremy R Coyle, Nima S Hejazi, Ivana Malenica, and Oleg Sofrygin.	*sl3: Pipelines for* *Machine Learning and Super Learning*, 2022. URL [https://github.com/tlverse/sl3.](https://github.com/tlverse/sl3) R package version 1.4.2.
[22] Nicholas T Williams and Ivan Diaz. lmtp: Non-parametric causal effects of feasible interventions based on modified treatment policies, 2020. https://github.com/nt-williams/lmtp[.](https://github.com/hoffmakl/lmtp_comprisks) R package version 1.1.1.5000.
